# Multi-modal single-cell and genetic integration defines cytotoxic T-cell regulatory states, HSPC suppression and inherited susceptibility in aplastic anaemia

**DOI:** 10.64898/2026.08.18.26360745

**Authors:** Fatimah M. Madkhaly, Menna Arafat

**Affiliations:** Department of Basic Medical Sciences (Pathology), Faculty of Medicine, Jazan University, Saudi Arabia; Mansoura University, Mansoura, Egypt

**Author notes:** Corresponding authors: Fatimah M. Madkhaly, Menna Arafat. These authors contributed equally to this work.

## Abstract

Acquired aplastic anaemia is caused by immune-mediated loss of haematopoietic stem and progenitor cells (HSPCs), but the regulatory states that sustain cytotoxic immunity and their relationship to inherited susceptibility remain incompletely understood. We integrated two single-cell RNA-sequencing cohorts spanning healthy, non-severe and severe aplastic anaemia with single-cell chromatin accessibility profiling, genome-wide association meta-analysis, Bayesian fine-mapping and stratified LD-score regression.

Single-cell transcriptomics revealed a coordinated shift across the immune and haematopoietic compartments. Cytotoxic CD8⁺ and γδ T cells converged on a shared NKG7/CCL5/PRF1 effector program, indicating that cytotoxic differentiation extends across T-cell lineages. Effector-memory T cells combined inflammatory signalling with SOCS, DUSP, TNFAIP3, RGS1 and TOX, consistent with sustained stimulation accompanied by extensive feedback regulation. With increasing disease severity, these inflammatory states were further coupled to hypoxic, oxidative and unfolded-protein-response programmes, suggesting qualitative remodeling of the immune compartment rather than uniform amplification of perforin-granzyme expression.

Single-cell chromatin accessibility provided a regulatory counterpart to these transcriptional states. Naive and memory-associated cells retained TCF7/LEF1/BACH2 accessibility, whereas cytotoxic cells acquired coordinated accessibility across CCL5, NKG7, PRF1, granzymes and killer-receptor loci. Pseudotime, motif activity and integrated RNA-chromatin profiles positioned AP-1, NFAT and TBX21 along this transition, linking loss of memory-associated regulation to acquisition of cytotoxic effector competence.

Genetic meta-analysis independently recovered association at the HLA-B region, reinforcing antigen presentation as the principal inherited susceptibility axis. Fine-mapping additionally prioritized a non-HLA locus without resolving its effector gene, while stratified LD-score regression found no detectable preferential enrichment of common-variant heritability within effector-memory or cytotoxic regulatory elements. Integrated with the cellular data, these findings support a mechanistic hierarchy in which HLA-linked antigen presentation establishes the selective context, persistent cytotoxic T-cell state remodeling maintains pathogenic immune pressure, and IFNγ-responsive HSPC suppression translates this pressure into haematopoietic failure.

## Introduction

Acquired aplastic anaemia is an immune-mediated bone-marrow-failure syndrome characterized by hypocellular marrow and peripheral cytopenias. In most acquired cases, activated cytotoxic lymphocytes target haematopoietic stem and progenitor cells (HSPCs), inflammatory cytokines suppress surviving progenitors, and clinical responses to immunosuppression provide functional support for this pathogenic model ^1,2^. The disorder is nevertheless heterogeneous. Variation in clinical severity, incomplete haematopoietic recovery, relapse and clonal evolution indicates that immune injury is not a single uniform state but a dynamic interaction among pathogenic lymphocytes, susceptible HSPCs and immune-escape clones.

Human genetic and clonal observations provide unusually direct evidence for antigen-selective pressure in aplastic anaemia. Copy-neutral 6p loss of heterozygosity and acquired HLA class I mutations can remove disease-associated HLA alleles from surviving haematopoietic clones, permitting escape from T-cell recognition ^3,4^. Germline association studies independently implicate the major histocompatibility complex, including HLA-DPB1 and HLA-B-linked variation^5^. High-resolution single-cell genomic reconstruction has now shown that several independent HLA-loss clones can coexist within the same patient, repeatedly converge on inactivation of HLA risk alleles and, in some cases, originate years before diagnosis ^6^. These findings place antigen presentation and immune-driven clonal selection at the centre of disease biology, but they do not define the transcriptional and epigenetic states through which antigen-experienced lymphocytes sustain marrow injury.

Single-cell studies have begun to resolve those states. Residual HSPCs from patients with aplastic anaemia show lineage-selective transcriptional disruption and persistent interactions with activated T cells ^7^. Functional studies have identified expanded T-cell clones capable of recognizing and eliminating HSPCs, including a molecular-mimicry mechanism linking viral and self-antigens ^8^.

Larger patient cohorts have further demonstrated convergence between NK cells and NK-like CD8+ effector populations, disease-associated T-cell-receptor signatures and treatment-responsive IFNG-IFNGR communication ^9,10^. Chromatin profiling in STAT1 gain-of-function-associated aplastic anaemia has also shown excessive STAT1 accessibility and pharmacologically reversible interferon signaling in a defined genetic context ^11^.

Important questions remain unresolved: how cytotoxic lymphocyte states scale with disease severity, which regulatory programs drive the transition from memory maintenance to effector differentiation, how these states converge on HSPC failure, and whether inherited susceptibility is concentrated within the same regulatory elements. We therefore integrated single-cell transcriptomic and chromatin-accessibility profiles with genome-wide association meta-analysis and statistical fine-mapping to resolve state-specific regulatory programs, connect transcription-factor activity with accessible chromatin and linked genes, and distinguish established HLA-mediated immune selection from non-HLA genetic signals that remain mechanistically unassigned.

## Results

### Integrated single-cell analysis identifies severity-associated cytotoxic lymphocyte programs in aplastic anaemia

Principal-component analysis of the two input datasets showed pronounced dataset-dependent separation before integration (Fig. 1A). Harmony correction markedly reduced this technical structure while preserving biological variation associated with disease status and severity (Fig. 1B). Azimuth mapping of the integrated atlas subsequently resolved the major lymphoid, myeloid, erythroid and progenitor compartments within a common transcriptional space (Fig. 1C).

**Fig. 1.**
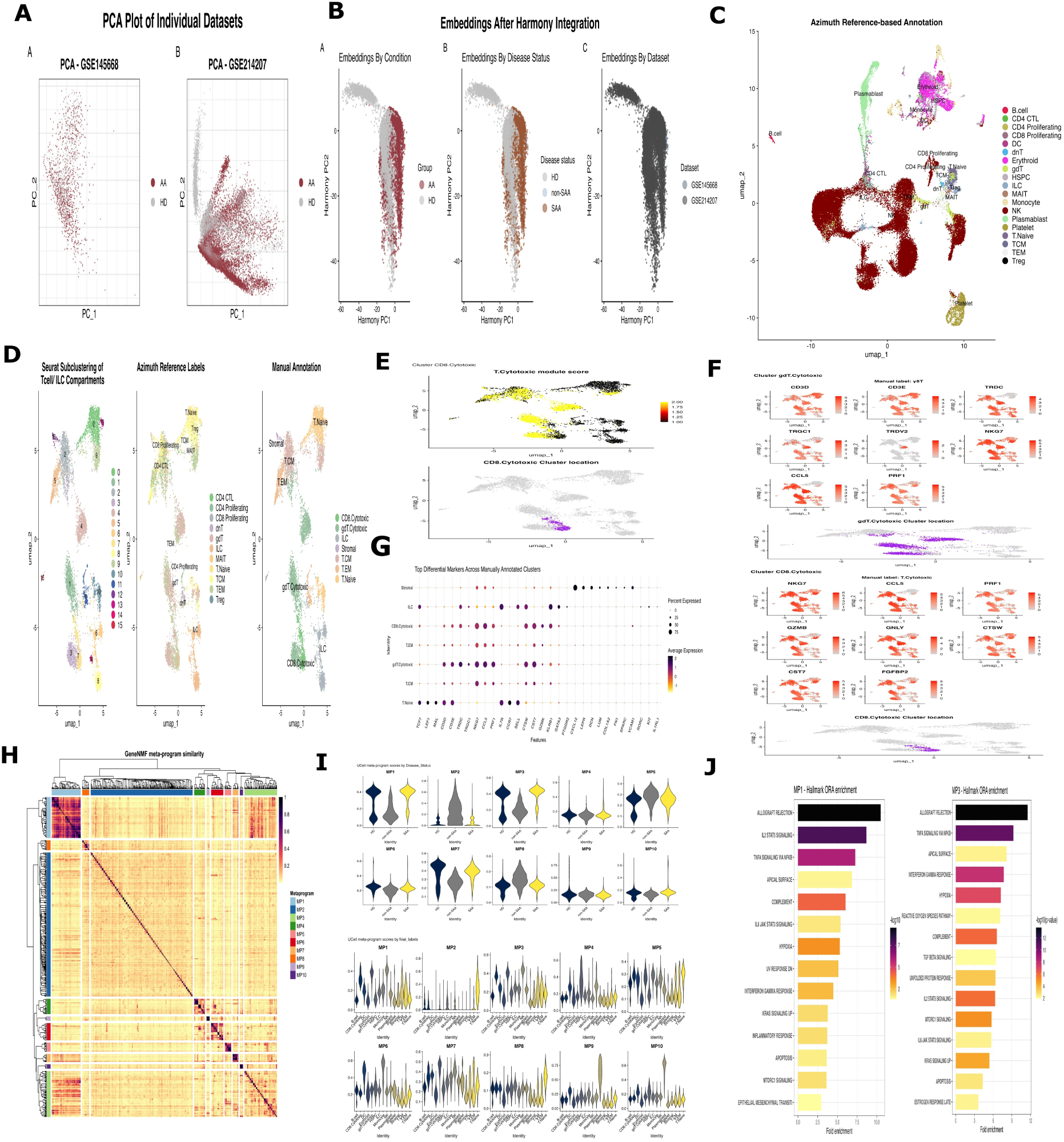
Integrated single-cell transcriptomic analysis identifies cytotoxic lymphocyte states in aplastic anaemia. **A)** Principal component analysis of the GSE145668 and GSE214207 datasets. **B)** Harmony-integrated embeddings coloured by condition, disease severity and dataset. **C)** UMAP of the integrated atlas annotated using Azimuth reference mapping. **D)** Subclustering of T-cell and innate lymphoid-cell populations, showing Seurat clusters, Azimuth labels and manually curated cell identities. **E)** Cytotoxicity-module scores and localisation of the cytotoxic CD8^+^ T-cell cluster. **F)** Expression of canonical lineage and effector genes supporting cytotoxic γδ T-cell and CD8^+^ T-cell annotation. **G)** Differentially expressed genes across manually annotated lymphocyte populations; dot size indicates the proportion of expressing cells and colour indicates average scaled expression. **H)** GeneNMF programme-similarity matrix defining ten consensus metaprogrammes (MP1-MP10). **I)** UCell scores for each metaprogramme stratified by disease status and lymphocyte identity. **J)** Hallmark pathway enrichment analysis of genes contributing to MP1 and MP3.

Re-clustering of T cells and innate lymphoid cells resolved discrete CD4+, CD8+, γδ T-cell, NK-cell and ILC states based on concordance among Seurat clusters, Azimuth labels and canonical lineage markers (Fig. 1D). Cytotoxicity scores localized sharply to cytotoxic lymphocyte compartments, with the curated cytotoxic CD8+ population occupying the highest-scoring region (Fig. 1E). These cells co-expressed NKG7, CCL5, PRF1, GZMB, GNLY, CTSW, CST7 and FGFBP2, whereas cytotoxic γδ T cells retained lineage-defining TRDC, TRGC1 and TRDV2 while acquiring the same core NKG7-CCL5-PRF1 effector module (Fig. 1F). Cluster-level marker analysis independently reproduced this separation from naive and non-cytotoxic lymphocyte states (Fig. 1G). The convergence of conventional CD8+ and γδ T cells on a common cytolytic program therefore reflects an effector state that crosses T-cell lineage boundaries rather than expansion of a single phenotypic compartment. This architecture closely parallels recent aplastic-anaemia studies identifying NK-like cytotoxic CD8+ populations, clonally expanded effector-memory T cells and dysregulated cytotoxic lymphocyte states ^9–11^.

GeneNMF further resolved ten consensus transcriptional metaprograms, MP1 to MP10, with distinct similarity structure (Fig. 1H). Their distribution was not explained by lymphocyte identity alone. MP1 and MP3 showed the clearest upward shift in severe aplastic anaemia (Fig. 1I), linking increasing disease severity to specific transcriptional states within the immune compartment. Because these distributions summarize individual cells, they define severity-associated shifts in cellular state rather than donor-level statistical effects.

MP1 was dominated by IFNγ, IFNα, TNF-NFκB, IL-2-STAT5, IL-6-JAK-STAT3, inflammatory-response and apoptosis signatures. MP3 retained this inflammatory architecture but added hypoxia, reactive-oxygen-species and unfolded-protein-response programs. Severe disease was therefore associated not simply with stronger cytotoxicity, but with coupling of inflammatory signaling to metabolic and proteostatic stress. This distinction is supported by recent experimental aplastic-anaemia data showing that oxidative metabolism can modulate T-cell phenotype and cytotoxic activity. In a 2026 study, eltrombopag reduced ROS in CD4+ and CD8+ T cells, altered T-cell composition and attenuated cytotoxic activity in experimental aplastic anaemia ^6^. The present MP3 signature therefore identifies oxidative stress as a plausible component of the severe-disease immune state, without establishing eltrombopag-responsive signaling or causality in the human samples analyzed here.

### Cell-type-resolved transcriptional programs link impaired haematopoiesis to immune activation

For cell-type-specific comparisons, genes with an absolute fold change of at least 1.5 and nominal *P* below 0.01 were highlighted in volcano plots, whereas inferential significance was defined by Benjamini-Hochberg adjusted *P* below 0.05. The most extensive disease-associated transcriptional disruption occurred in HSPCs (Fig. 2A). Severe aplastic anaemia showed coordinated loss of erythroid specification and maturation genes, including KLF1, GATA1, GFI1B, HBA1, HBA2, HBB, AHSP, GYPA, RHAG, ANK1 and SPTA1, together with suppression of haem-biosynthetic genes TMEM14C, FECH, UROD, ALAD and CPOX. Reduced CCNB1, CCNB2, CENPF, CDC20, PRC1 and UBE2C simultaneously marked depletion of proliferative progenitor activity (Fig. 2A, upper).

**Fig. 2.**
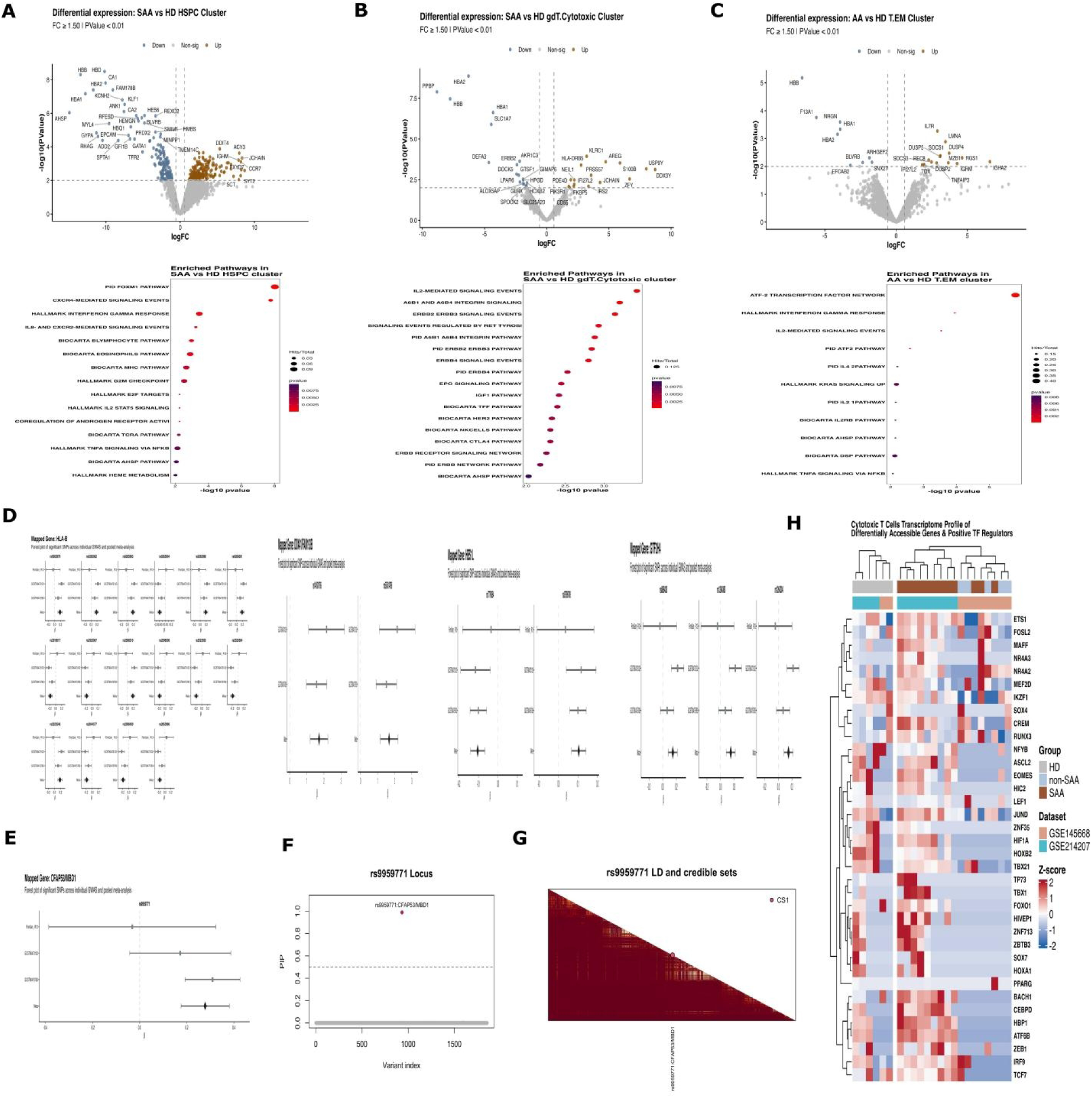
Cell-type-specific transcriptional dysregulation and genetic prioritization in aplastic anaemia. **A-C)** Volcano plots showing differentially expressed genes between severe aplastic anaemia (SAA) and healthy donor (HD) cells in the HSPC, γδ/cytotoxic T-cell and effector-memory T-cell (T_EM) clusters, respectively. Corresponding pathway-enrichment analyses are shown below each volcano plot. **D)** Forest plots showing cohort-specific and pooled effect estimates for SNPs significant in the METAL meta-analysis. **E)** Forest plot of the variant prioritized by both METAL meta-analysis and SuSiE fine-mapping. **F)** Posterior inclusion probabilities (PIP) across variants at the prioritized locus. **G)** Linkage-disequilibrium matrix for the corresponding locus. **H)** Heatmap of differentially accessible genes in cytotoxic T cells across HD, non-SAA and SAA samples; double asterisks indicate genes overlapping METAL-significant GWAS signals and scATAC-seq differentially accessible genes.

This loss of erythroid and cell-cycle output was accompanied by induction of IFI27L2, IFNGR1, SOCS1, HLA-DRB1, HLA-DRB5, HLA-DQB1, NFKB1, MAP3K8, GADD45B, DDIT4, RGS1, RGS2, RUNX3 and TOX, defining an HSPC state exposed to interferon, inflammatory and cellular-stress signals. Pathway enrichment reproduced the same reciprocal structure, with inflammatory cytokine programs rising as haematopoietic programs contracted (Fig. 2A, lower). The HSPC phenotype therefore connects immune activation to the cellular consequence that defines aplastic anaemia: loss of productive lineage output. This interpretation is supported by experimental evidence that sustained IFNγ disrupts HSPC composition and lineage differentiation, by IFNγ-dependent HSC loss mediated through the marrow microenvironment, and by single-cell evidence of selective lineage disruption in residual aplastic-anaemia HSPCs ^7,12,13^.

The γδ/cytotoxic T-cell compartment showed a narrower and mechanistically distinct disease-associated shift (Fig. 2B). Increased KLRC1, HLA-DRB5, IFI27L2, AREG, IRS2, PIK3R1, FKBP5, PDE4D and CD55 combined inhibitory-receptor, interferon-response and growth-factor signaling features, while enrichment mapped predominantly to IL-2, integrin and PI3K-AKT-associated pathways (Fig. 2B, lower). In parallel, reduced GIMAP6, LPAR6, HPGD, ALOX5AP, GLRX and SLC25A20 indicated altered lymphocyte homeostasis, lipid signaling and redox-metabolic regulation.

Importantly, PRF1, GZMB and related cytolytic genes were not coordinately increased in this disease comparison. Disease-associated remodeling of γδ T cells therefore involved signaling and adaptive-state changes rather than uniform amplification of their cytotoxic machinery. This pattern is distinct from the IL-17A-producing γδ T-cell phenotype described in a T-cell-activated subset of severe aplastic anaemia, where γδT17 frequency was linked to immune activation and disease severity ^14^.

Effector-memory T cells exhibited a complementary inflammatory state marked by strong intracellular feedback control (Fig. 2C). IL7R co-occurred with SOCS1, SOCS3, DUSP2, DUSP4, DUSP5, TNFAIP3 and RGS1, placing cytokine, MAPK, NFκB and G-protein signaling under multiple negative-feedback circuits. IFI27L2 and TOX further indicated sustained interferon exposure and adaptation to repeated stimulation. Pathway enrichment for IFNγ, IL-6, IL-1, IL-12, TNF-NFκB, ATF2 and RAS-KRAS signaling supported the same configuration (Fig. 2C, lower). The resulting state is therefore better defined as chronically stimulated and signal-adapted than as acutely activated or uniformly exhausted. This effector-memory phenotype provides the lymphocyte counterpart to the HSPC response. Aplastic anaemia has long been associated with expansion of CD8+ effector-memory cells and restricted TCR repertoires, while recent single-cell analysis shows that severe disease is dominated by clonally expanded effector-memory populations with sustained cytotoxic programs and IFNγ-centered communication with haematopoietic cells ^10,15,16^.

Across compartments, the data therefore resolve a linked disease architecture in which antigen- experienced cytotoxic lymphocytes acquire inflammatory and stress-adapted states as residual HSPCs simultaneously lose erythroid and proliferative programs and acquire interferon-responsive stress signaling.

### Genetic fine-mapping and chromatin integration implicate antigen presentation and cytotoxic T-cell state regulation

Cross-cohort meta-analysis identified concordant association signals at the HLA-B region, with consistent cohort-specific and pooled effects for the significant variants (Fig. 2D). This germline association falls within the same antigen-presentation axis that undergoes recurrent somatic selection in aplastic anaemia. HSPC clones carrying copy-neutral 6p loss of heterozygosity or somatic HLA class I loss-of-function mutations preferentially eliminate specific HLA alleles, providing a selective advantage under HLA-restricted immune pressure ^3,4,17^. High-resolution single-cell genomics has further shown that multiple independent HLA-loss clones can coexist in the same patient, repeatedly converge on inactivation of HLA risk alleles and arise years before diagnosis ^18^. The HLA signal in Fig. 2D therefore reinforces an established selective axis of aplastic anaemia rather than defining a new mechanism.

Outside the HLA region, rs9959771 was supported by the METAL meta-analysis and retained the highest statistical support after SuSiE fine-mapping. Its cohort-specific and pooled effect is shown in Fig. 2E, posterior inclusion probability concentrates on rs9959771 in Fig. 2F, and the local linkage- disequilibrium structure defines the corresponding credible interval in Fig. 2G. Nearest-gene annotation mapped this signal to MBD1, but the genetic evidence does not establish MBD1 as the effector gene. MBD1 is a methyl-CpG-binding transcriptional repressor that binds methylated CpGs and recruits repressive chromatin machinery ^19,20^. Recent functional work in myelodysplastic neoplasms showed that an aberrant long MBD1 isoform suppresses HSPC cycling, impairs terminal erythroid differentiation and reduces haematopoietic reconstitution under increased demand ^21^.

MBD1 can also regulate methylation-dependent responsiveness to IFNγ in a non-haematopoietic experimental system ^22^. The present data therefore prioritize rs9959771 as a candidate regulatory locus, while assignment of MBD1 requires colocalizing eQTL or caQTL evidence, physical enhancer-promoter linkage or allele-specific perturbation.

### Chromatin-linked cytotoxic T-cell states converge on the IFNγ injury axis

Integration of differential chromatin accessibility with predicted transcription-factor regulation resolved two opposing programs within cytotoxic T cells (Fig. 2H). ETS1, FOSL2, RUNX3 and EOMES defined an effector-associated state, whereas TCF7, LEF1, FOXO1 and IKZF1 marked a less-differentiated, memory-associated state ^26^. IRF9, HIF1A, ATF6B, CEBPD, ZEB1 and BACH1 added interferon, metabolic and stress-responsive variation. Genes marked by double asterisks additionally satisfied the analysis-specific overlap between genetic association and differential chromatin accessibility.

The disease-relevant feature was the effector pole. In aplastic anaemia, T-bet is increased in T cells and directly promotes IFNG transcription ^23,24^, while experimental marrow failure shows that IFNγ increases pro-apoptotic signaling in HSPCs, impairs repopulating capacity and increases susceptibility to immune-mediated elimination ^25^. Clinical status only partly explained this structure, with clustering also influenced by dataset origin, arguing against a uniform SAA-specific transcriptional state.

### Single-cell chromatin accessibility resolves homeostatic and cytotoxic T-cell regulatory programs

Unsupervised scATAC-seq resolved naive, central-memory, effector-memory, cytotoxic, MAIT and Th17-like T-cell states (Fig. 3A). GeneScoreMatrix patterns, marker-gene activity and locus-level accessibility independently reproduced these annotations (Fig. 3B–D) and exposed a dominant regulatory contrast. Naive and central-memory cells retained accessibility at IL7R, CCR7, SELL, MAL, LRRN3, TCF7, LEF1 and BACH2, whereas cytotoxic cells opened a coordinated CCL5-NKG7- PRF1-CTSW-GZMH-GZMM program together with KLRD1, KLRC2, KLRC3, FCGR3A, ADGRG1 and S1PR5. Effector-memory cells occupied the intervening state, retaining cytotoxic accessibility while acquiring regulatory loci including CTLA4, TIGIT, TNFRSF1B, NDFIP1 and ZFP36. This opposition between TCF7-LEF1-BACH2 and cytotoxic chromatin is consistent with the established role of TCF1/LEF1 and BACH2 in preserving memory potential and restraining activation-responsive effector programs ^28,29^.

**Fig. 3.**
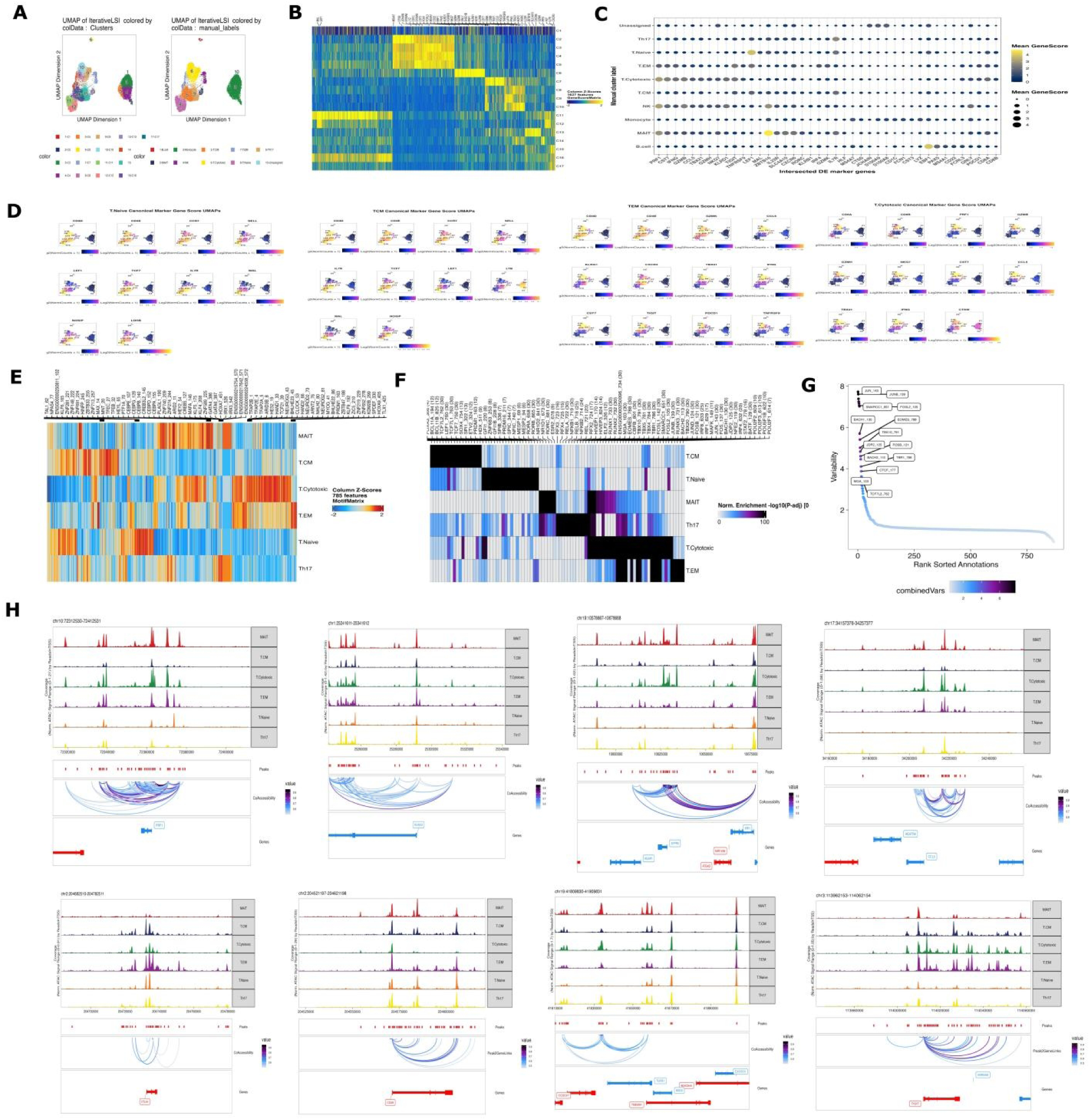
Single-cell chromatin accessibility delineates T-cell states and regulatory programs in aplastic anaemia. **A)** UMAP of scATAC-seq profiles coloured by unsupervised cluster and annotated T-cell state. **B)** Heatmap of cluster-specific gene-activity scores. **C)** Dot plot showing marker gene activity across annotated populations. **D)** UMAP feature plots of canonical lineage and state markers. **E)** Heatmap of transcription-factor motif accessibility across T-cell subsets. **F)** Enrichment of transcription-factor motifs in cell-state-specific accessible regions. **G)** Rank-ordered variability of motif activity, with highly variable motifs labelled. **H)** Representative pseudobulk accessibility tracks showing peaks, co-accessibility links and peak-to-gene associations across T-cell states.

Motif analysis independently resolved the same regulatory architecture (Fig. 3E–G). TCF7 and LEF1 were associated with less-differentiated states, whereas TBX21, EOMES, ETS1, JUND, FOSL2, IRF9, HIF1A and ZEB1 characterized activated and cytotoxic states. Enrichment of T-box, RUNX, ETS, AP-1, BATF and IRF motifs is consistent with established regulators of CD8+ effector differentiation and activation-induced chromatin remodeling ^30–32^. The relevance to aplastic anaemia is particularly direct for TBX21, because increased T-bet in patient T cells drives enhanced IFNG transcription ^23^.

Peak-to-gene correlation and co-accessibility provided an independent structural basis for this distinction (Fig. 3H). The cytotoxic links included CCL5, PRF1, NKG7, granzymes, KLR genes, S1PR5, RUNX3, ZEB2 and NFIL3. Notably, S1PR5 provides a direct disease-specific anchor, as activated S1PR5+ CD8+ T cells and NK cells are expanded in severe aplastic anaemia ^10^, while the broader CCL5-NKG7-PRF1-granzyme module recapitulates the NK-like and clonally expanded cytotoxic states independently identified in aplastic anaemia ^9,10^. Thus, two complementary chromatin-linking approaches converge on an AA-relevant cytotoxic regulatory program, strengthening its biological assignment without implying causal enhancer-gene relationships.

### Pseudotime resolves a regulatory transition from memory maintenance to cytotoxic effector function

ArchR pseudotime ordered naive and central-memory T cells through an effector-memory intermediate toward the cytotoxic state (Fig. 4A, B), consistent with the antigen-experienced effector expansions reported in aplastic anaemia ^8–10,15^. Across chromatin accessibility, motif activity and integrated RNA-scATAC profiles, this transition progressed from IL7R-LEF1-BACH2 memory- associated regulation through AP-1/NFAT/TBX21 activation to a terminal CCL5-FGFBP2-NKG7- GZMB-PRF1 cytotoxic program (Fig. 4C, D). This coordinated ordering supports progressive regulatory remodeling from memory maintenance to effector competence rather than isolated activation of cytotoxic genes ^29,33–36^. The concordance across accessibility, motif activity and integrated expression indicates an organized regulatory transition rather than isolated induction of individual cytotoxic genes.

**Fig. 4.**
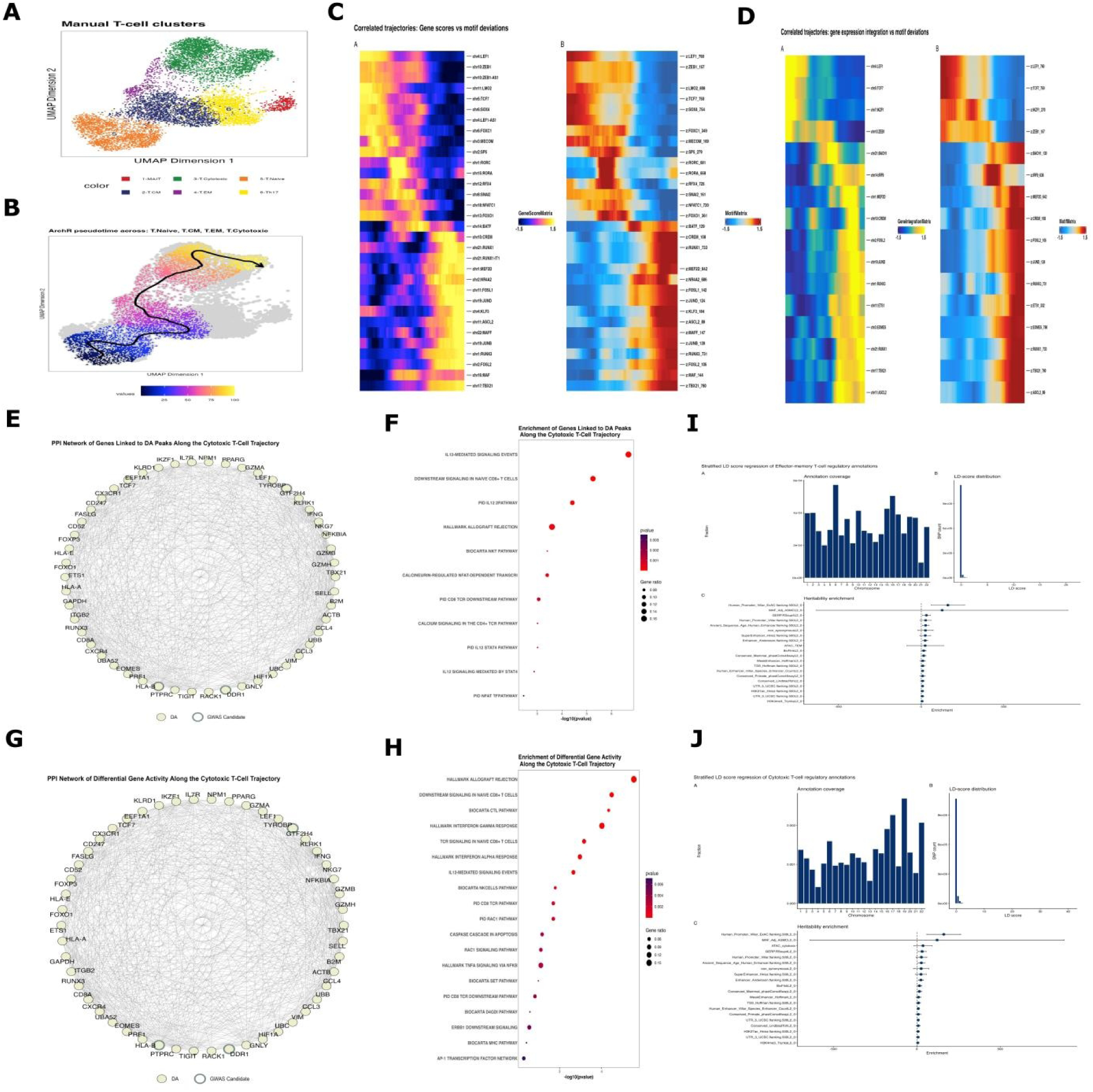
Pseudotime analysis defines regulatory and genetic programs across T-cell differentiation in aplastic anaemia. **A)** UMAP of manually annotated T-cell states. B) ArchR pseudotime trajectory from naive and central-memory to effector-memory and cytotoxic T cells. **C)** Correlated pseudotime patterns of scATAC-seq gene scores and transcription-factor motif deviations. **D)** Correlated pseudotime patterns of integrated gene activity and motif deviations. **E)** Protein-protein interaction network of genes linked to differentially accessible peaks along the cytotoxic T-cell trajectory. **F)** Pathway enrichment of genes linked to these peaks. **G)** Protein-protein interaction network of genes with differential activity along the cytotoxic trajectory. H) Pathway enrichment of differentially active genes. **I, J)** Stratified LD score regression of effector­memory and cytotoxic T-cell trajectory annotations, respectively, showing annotation coverage, LD-score distributions and heritability enrichment. Gene activity represents the scRNA-scATAC GenelntegrationMatrix, whereas peak accessibility is derived from the scATAC-seq PeakMatrix.

Trajectory-linked accessibility and integrated gene activity independently converged on TCR, IL-12- STAT4, NFAT-AP-1, IFNγ, TNF-NFκB and FAS signaling (Fig. 4E–H), connecting receptor-driven activation and type 1 polarization to perforin-granzyme and death-receptor effector mechanisms.

This regulatory sequence is directly relevant to aplastic anaemia, where IFNγ-producing and perforin-positive T cells are increased in active disease, IFNγ impairs HSPC function, and clonally expanded antigen-reactive T cells can eliminate haematopoietic progenitors ^8,12,37,38^.

Stratified LD-score regression did not support preferential localization of common-variant heritability within either effector-memory or cytotoxic trajectory regulatory elements (Fig. 4I, J). Across traits, enrichment estimates clustered around the null. This absence of detectable enrichment is consistent with a genetic architecture dominated by HLA-mediated antigen-presentation effects and acquired clonal selection, rather than broad non-HLA polygenic enrichment across cytotoxic T-cell regulatory elements ^3–5,8–10^.

## Discussion

The integrated data support a disease model in which persistent antigen-driven cytotoxic immunity imposes selective pressure on HSPCs while simultaneously disrupting haematopoietic output. In severe aplastic anaemia, residual HSPCs lose erythroid, haem-biosynthetic and proliferative programs and acquire interferon-responsive and stress-adaptive states. Cytotoxic CD8+ and γδ T cells converge on a shared effector phenotype, but disease-associated variation extends beyond perforin and granzyme expression to sustained receptor and cytokine signaling accompanied by metabolic adaptation and induction of intracellular feedback regulators that constrain JAK/STAT, MAPK and NF-κB signaling. The HSPC and lymphocyte phenotypes converge most clearly on IFNγ signaling. IFNγ disrupts HSPC composition and lineage differentiation in experimental aplastic anaemia ^12^, and ingle-cell studies identify enhanced IFNG-IFNGR communication between cytotoxic lymphocytes and haematopoietic cells ^10^.

Our data resolve the corresponding target-cell response as coordinated lineage suppression, proliferative failure and cellular stress. In parallel, induction of SOCS, DUSP, TNFAIP3 and RGS regulators within effector-memory and cytotoxic populations indicates adaptation to sustained signaling rather than transient activation. This interpretation is consistent with oligoclonal effector- memory expansion, disease-associated TCR signatures and persistent clonal remodeling after immunosuppression ^9,10,15^.

The chromatin landscape provides a regulatory basis for this persistence. TCF7, LEF1, BACH2 and FOXO1 define a memory-maintaining program that gives way along pseudotime to AP- 1/NFAT/TBX21 activation and acquisition of cytotoxic effector machinery. This ordering is consistent with BACH2-mediated restriction of activation-responsive enhancers, AP-1-dependent chromatin opening and T-bet-driven effector differentiation ^29, 35, 36^.

Importantly, T-bet is increased in aplastic-anaemia T cells and directly promotes IFNG transcription^23^. The inferred trajectory therefore links persistence and pathogenicity within the same antigen- experienced compartment: memory-associated regulatory capacity may sustain the population, while progressive chromatin remodeling enables acquisition of IFNγ and cytotoxic effector function.

The HLA findings provide the strongest disease-specific evidence in the study. Germline HLA association, copy-neutral 6p loss of heterozygosity and somatic HLA class I mutations converge on HLA-restricted antigen presentation as the dominant disease-specific genetic axis. The 2026 single- cell genomic study adds an evolutionary dimension by showing a median of multiple HLA-loss events per patient, recurrent convergence on HLA risk-allele inactivation, origins years before diagnosis and enrichment of long-lived clones in the CD34+ HSPC compartment ^18^. These observations support a model in which immune pressure removes susceptible HSPCs while favoring clones that reduce antigen presentation. HLA loss is therefore not simply a marker of clonality, but evidence of the selective pressure exerted by the pathogenic immune response ^8^.

The non-HLA findings are substantially less definitive. rs9959771 is statistically prioritized, and MBD1 is biologically plausible given its role in methylation-dependent transcriptional repression and its reported effects on HSPC cycling and erythroid maturation ^19–21^, but neither proximity nor fine- mapping establishes MBD1 as the effector gene. Consistent with this uncertainty, stratified LD-score regression showed no preferential enrichment of common-variant heritability within effector-memory or cytotoxic regulatory elements. The pronounced cytotoxic chromatin phenotype therefore appears more consistent with an acquired immune state shaped by antigenic selection than with a broadly distributed non-HLA polygenic architecture.

These findings remain bounded by the constraints of cohort composition and assay resolution. Residual dataset structure after integration and the limited scATAC-seq cohort, including a STAT1 gain-of-function-associated case, restrict claims of universal disease specificity. Moreover, peak-to- gene links, co-accessibility and motif correlations identify regulatory associations rather than causal enhancer activity or transcription-factor occupancy. The next priority is therefore longitudinal, clone-resolved multiomic analysis before and after immunosuppression, integrating scRNA-seq, scATAC- seq, paired TCR sequencing and HLA genotyping to determine whether antigen-experienced T-cell clones progress along the inferred regulatory trajectory in parallel with expansion of HLA-loss HSPCs. Functional perturbation of TBX21, BACH2 and AP-1/NFAT, together with allele-specific testing of fine-mapped non-HLA variants, will be required to distinguish regulatory signatures of persistent activation from mechanisms that directly drive HSPC injury, immune escape and clonal persistence.

## Conclusion

Aplastic anaemia appears to involve a coordinated shift in both immune regulation and haematopoietic function. Cytotoxic CD8⁺ and γδ T cells converge on a shared effector program, while chromatin accessibility places this phenotype along a transition from TCF7/LEF1/BACH2 memory-associated regulation to AP-1/NFAT/TBX21 activation and cytotoxic effector competence. In parallel, residual HSPCs lose erythroid and proliferative programs and acquire interferon-responsive stress states, linking immune dysregulation to impaired haematopoietic output.

The association of severe disease with oxidative, hypoxic and proteostatic stress suggests that progression is accompanied by a broader reorganization of the immune state rather than by increased cytotoxic gene expression alone. Recovery of the HLA-B susceptibility signal places this cellular phenotype within an antigen-presentation background that may shape which immune responses are selected and maintained.

Taken together, the study separates three features that are often treated as a single process: genetic susceptibility, persistence of cytotoxic immune states, and loss of HSPC function. Their separation may help explain why immune activity, disease severity and haematopoietic recovery do not necessarily change in parallel, and points to persistence of the immune state rather than cytotoxic magnitude alone as a critical feature of aplastic-anaemia biology.

## Methods

### Data acquisition and study design

Processed single-cell RNA-sequencing count matrices and sample metadata were obtained from GEO accessions GSE145668 and GSE214207. GSE145668 is the 3′ RNA-sequencing SubSeries within the GSE145669 SuperSeries and contains HSPCs and T cells from bone marrow and peripheral blood of patients with aplastic anaemia and healthy donors ^7^. GSE214207 contains bone- marrow and peripheral-blood NK-cell profiles from patients with severe aplastic anaemia and healthy donors ^39^. Single-cell chromatin-accessibility fragment files and associated metadata were obtained from Zenodo record 5747633, derived from the study of STAT1 gain-of-function-associated aplastic anaemia ^11^. Sample, donor, tissue, disease status, dataset and treatment identifiers were retained throughout the analysis.

Genome-wide association summary statistics were obtained from FinnGen release 13 and the NHGRI-EBI GWAS Catalog studies GCST90473132 and GCST90475783 ^40–42^. These GWAS datasets were previously analyzed by Madkhaly and Arafat (2026). In the present study, the same summary-statistic datasets were re-used for additional downstream analyses, including fine-mapping and integration with single-cell transcriptomic and chromatin-accessibility data.

### Single-cell RNA-sequencing analysis

Count matrices from GSE145668 and GSE214207 were imported into Seurat ^43^and restricted to genes shared between the two datasets. Cells from peripheral blood were retained for the combined analysis, and the proportion of mitochondrial transcripts was calculated for each cell. Quality-control filtering retained cells with more than 200 and fewer than 10,000 detected RNA features, and mitochondrial RNA content below 30%. Doublets were identified using scDblFinder with sample identity supplied as the grouping variable, and only cells classified as singlets were retained.

Mitochondrial and ribosomal genes matching the prefixes MT-, RPS or RPL were removed after cell-level filtering. After quality-control filtering, 62,072 cells from 22 samples were retained, comprising 45,597 cells from patients with aplastic anaemia and 16,475 cells from healthy donors. Among the aplastic-anaemia cells, 44,870 were derived from patients with severe aplastic anaemia and 727 from patients with non-severe aplastic anaemia.

The transcriptomic datasets were integrated in principal-component space using Harmony ^44^, with dataset and sample identity modeled as technical covariates. Disease severity, tissue and cell identity were not regressed because they were biological variables of interest. A shared-nearest- neighbor graph was constructed from the Harmony dimensions and used for graph-based clustering and UMAP visualization. Integration was assessed by examining the distribution of datasets, samples and biological groups in the common embedding.

Initial cell identities were assigned by mapping the integrated object to the Azimuth human PBMC reference and were reviewed using canonical lineage markers and cluster-specific expression ^45^. T- cell and innate-lymphoid populations were subset and reanalyzed. Naive, central-memory, effector- memory, cytotoxic, regulatory, γδ T-cell states were assigned using transferred labels and manually-labelled with established marker combinations. NK cells were distinguished by KLRD1, FCER1G, TYROBP, NKG7 and cytotoxic effector genes in the absence of a conventional T-cell-receptor program. Cytotoxic activity was quantified using UCell with a predefined signature containing GNLY, NKG7, PRF1, CTSW, CCL5, FGFBP2 and granzyme genes ^46^. Cluster markers were identified using two-sided Wilcoxon rank-sum tests.

Recurrent transcriptional programs were derived using GeneNMF non-negative matrix factorization. Factorization was performed across multiple ranks and samples, programs with similar gene-weight profiles were grouped into consensus metaprograms, and ten stable programs (MP1-MP10) were retained ^47^. program activity was quantified in individual cells using UCell and summarized across disease groups and lymphocyte identities. Genes contributing to MP1 and MP3 were tested for enrichment in Molecular Signatures Database Hallmark pathways ^48^.

### Differential expression and pathway analysis

Disease-associated expression was evaluated separately in HSPCs, γδ/cytotoxic T cells, effector- memory T cells and NK cells. Donor was treated as the biological replicate by aggregating raw counts within each donor and cell identity before model fitting. Low-abundance genes were removed, library sizes were normalised and disease-associated expression was tested with negative-binomial models implemented using edgeR ^49^. Dataset and tissue were included as covariates where supported by the design. Genes with absolute fold change of at least 1.5 and nominal P below 0.01 were highlighted in volcano plots and considered differentially expressed (DE). Differentially expressed genes were tested for over-representation in Hallmark, PID and BioCarta pathways using the genes retained in each cell-type-specific model as the background. Enrichment was evaluated using a hypergeometric test and corrected for multiple testing with clusterProfiler-compatible procedures ^50^.

### Single-cell chromatin-accessibility analysis

Single-cell ATAC-sequencing fragment files from three untreated aplastic anaemia samples were processed using ArchR [51]. Sample-specific Arrow files were generated, predicted doublets were removed, and genome-wide chromatin accessibility was quantified using a 500-bp TileMatrix. The initial scATAC-seq dataset contained 15,157 cells, with a median transcription start site enrichment score of 14.48 and a median of 7,585 unique fragments per cell. Iterative latent semantic indexing was performed on the TileMatrix, followed by Harmony correction to reduce sample-associated technical variation. UMAP embedding and graph-based clustering were then calculated from the corrected dimensions.

Gene activity was quantified using the ArchR GeneScoreMatrix. Cell clusters were manually annotated based on canonical lineage markers, differential GeneScore markers and overlap with predefined marker-gene sets. Naive and central-memory T cells were identified by markers including CD3D, CD3E, CCR7, SELL, TCF7 and LEF1; effector-memory T cells by GZMK, CCL5, KLRG1, TBX21 and IFNG; cytotoxic T cells by CD8A, CD8B, PRF1, GZMB, GZMH, NKG7 and CTSW; and MAIT cells by SLC4A10, KLRB1, ZBTB16, CXCR6 and TRAV1-2. NK-cell annotation was supported by KLRD1, GNLY, NKG7, PRF1, GZMB, FCGR3A and TYROBP.

Pseudobulk chromatin-accessibility profiles were generated for each manually annotated T-cell state, followed by MACS2 peak calling and construction of a reproducible PeakMatrix ^52^. Differentially accessible peaks were identified using Wilcoxon tests with transcription-start-site enrichment and log10 fragment number included as bias covariates. Marker peaks were retained at FDR ≤ 0.1 and Log2FC ≥ 0.5. Motif enrichment was assessed within these marker peaks, and chromVAR was used to quantify motif accessibility across T-cell states. Differential chromVAR motifs were retained at FDR ≤ 0.1 and mean deviation difference ≥ 0.1.

Transcription-factor motif annotations were added to the peak set using CIS-BP position-weight matrices ^53^. Motif enrichment was tested within cell-state-specific accessible peaks, while chromVAR was used to calculate bias-corrected motif deviation scores and identify motifs with differential accessibility across T-cell states ^54^. Positive transcription-factor regulators were identified by correlating integrated scRNA-seq expression from the GeneIntegrationMatrix with motif accessibility in the MotifMatrix. Regulators were retained when the correlation was > 0.5, adjusted P < 0.01, and the motif-accessibility change exceeded the 75th percentile of the observed distribution.

Peak-to-gene links were inferred by correlating chromatin accessibility with integrated scRNA-seq expression from the GeneIntegrationMatrix using IterativeLSI dimensions. Links were retained at a correlation cutoff of ≥0.45 and visualized at FDR ≤1 × 10⁻⁴ . Peak co-accessibility was calculated independently from scATAC-seq accessibility data using a correlation cutoff of ≥0.50.

Representative browser tracks were generated across manually annotated T-cell states to display chromatin accessibility together with peak-to-gene and co-accessibility links.

### Trajectory and network analysis

A supervised ArchR trajectory was constructed across naive, central-memory, effector-memory and cytotoxic T-cell states using IterativeLSI dimensions. Gene activity, peak accessibility, chromVAR motif deviations and integrated scRNA-seq expression were profiled along pseudotime. Dynamic features were identified using matrix-specific variance thresholds, and coordinated gene-motif changes were assessed using trajectory correlation analysis. Correlated features were retained at |r| ≥0.50 and FDR ≤0.05. Genes showing differential gene activity along the cytotoxic T-cell trajectory and genes linked to differentially accessible trajectory peaks were analyzed separately. Protein- protein interaction networks were constructed using STRING v12.0 interactions, retaining interactions with a combined score >400 ^55^. Duplicate edges and self-loops were removed, and networks containing more than 50 nodes were reduced while prioritizing genetically supported candidate genes. Functional enrichment of the two trajectory-derived gene sets was performed using clusterProfiler against selected Bader Lab gene-set collections, including MSigDB, Hallmark, NCI and BioCarta pathways.

### GWAS meta-analysis and fine-mapping

Aplastic-anaemia summary statistics from FinnGen release 13 and GWAS Catalog studies GCST90473132 and GCST90475783 were harmonized to a common genome assembly, effect allele and reference allele. Duplicate records, unresolved allele mismatches, ambiguous variants and variants lacking effect estimates or standard errors were removed. Cohort-specific effects were combined using inverse-variance-weighted fixed-effect meta-analysis in METAL ^56^. Genome-wide significance was defined as P below 5 x 10^-8^.

Associated loci were fine-mapped with SuSiE-RSS using an ancestry-matched linkage- disequilibrium reference ^57,58^. Posterior inclusion probabilities were calculated for all variants and 95% credible sets were derived from the fitted single-effect components. Linkage disequilibrium and posterior inclusion probabilities were visualized using a common variant order.

### Integration of genetic and single-cell evidence

Genome-wide significant and fine-mapped variants were integrated with scATAC-seq regulatory features. Genes supported by genetic evidence together with differential accessibility or positive transcription-factor-regulator signals in cytotoxic T cells were then evaluated in the scRNA-seq atlas. Cytotoxic T-cell expression was aggregated at the sample level, standardized to gene-wise Z scores, and hierarchically clustered using Spearman distance. Genetically supported candidate genes were highlighted in the resulting heatmap.

Cell-state-specific annotations for stratified linkage-disequilibrium score regression were generated from differentially accessible peaks in cytotoxic and effector-memory T cells relative to naive T cells. For S-LDSC, open-chromatin peaks were retained at FDR ≤0.05 and Log2FC ≥1, lifted from hg19 to GRCh38, and restricted to autosomes. S-LDSC was performed separately for cytotoxic and effector- memory T-cell annotations using baselineLD v2.2, HapMap3 SNPs excluding the MHC region, and European 1000 Genomes Phase 3 LD reference data ^59,60^.

### Statistical analysis and reproducibility

All statistical tests were two sided unless otherwise stated. Multiple testing was controlled using the Benjamini-Hochberg procedure. Donors were considered biological replicates for disease comparisons; individual cells were used for clustering, annotation, signature scoring and descriptive marker analysis.

## Data availability

The 3’ single-cell RNA-sequencing data analyzed from the Zhu et al. study are available under GEO accession GSE145668, a SubSeries of GSE145669 ^7^. The NK-cell single-cell RNA-sequencing data are available under GSE214207 ^39^. Single-cell ATAC-sequencing data and associated code are available from Zenodo record 5747633 ^11^. GWAS summary statistics were obtained from [FinnGen release 13](https://r13.finngen.fi/) and the NHGRI-EBI GWAS Catalog studies GCST90473132 and GCST90475783 ^40–42^.

## Author Contributions

Fatimah M. Madkhaly: supervision, manuscript review, editing, and writing. Menna Arafat: Data curation, formal analysis, visualization, and writing.

---

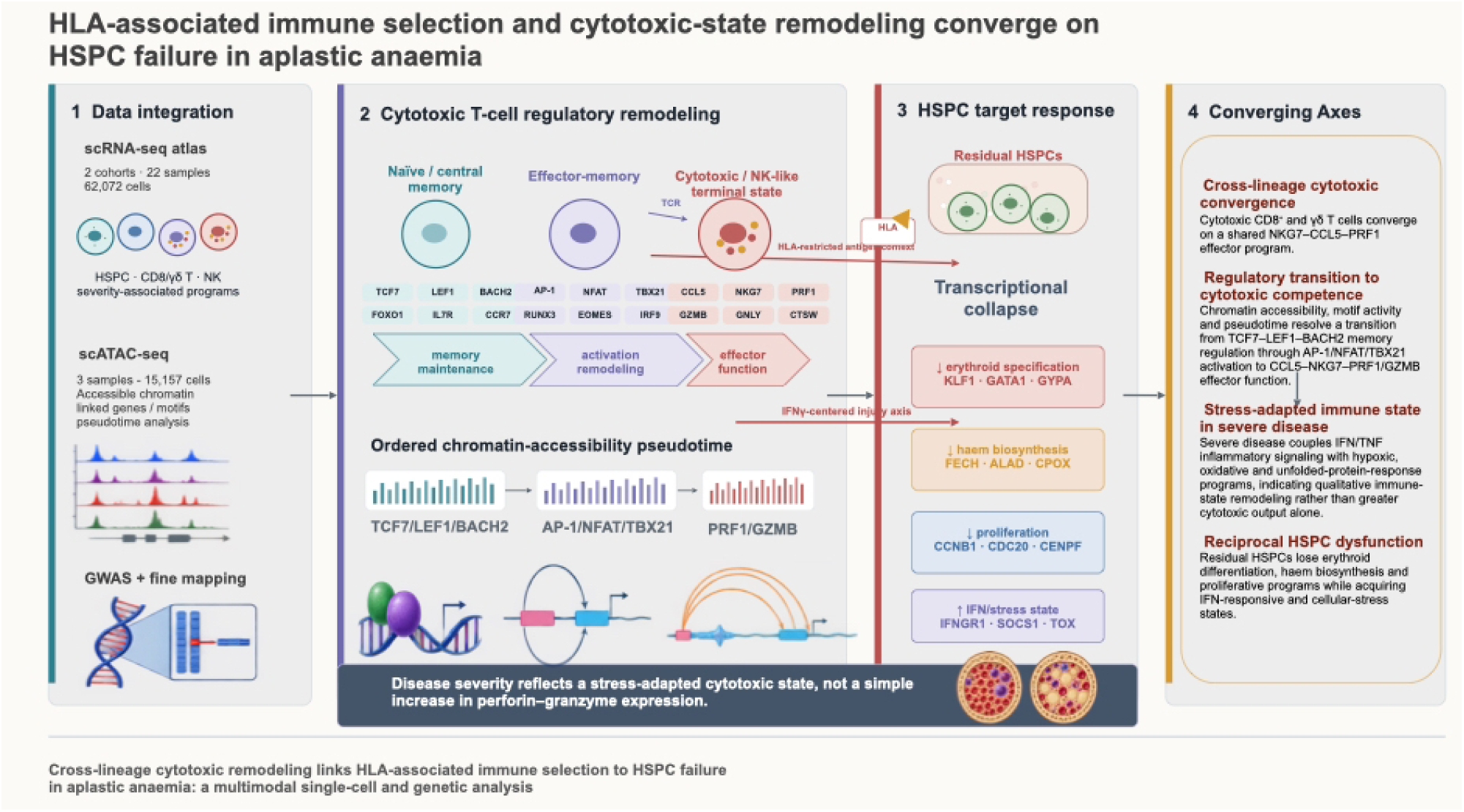

## References

1. Young NS, Calado RT, Scheinberg P. Current concepts in the pathophysiology and treatment of aplastic anemia. Blood. 2006;108(8):2509–2519. doi:10.1182/blood-2006-03-010777

2. Luzzatto L, Risitano AM. Advances in understanding the pathogenesis of acquired aplastic anaemia. Br J Haematol. 2018;182(6):758–776. doi:10.1111/bjh.15443

3. Katagiri T, Sato-Otsubo A, Kashiwase K, et al. Frequent loss of HLA alleles associated with copy number-neutral 6pLOH in acquired aplastic anemia. Blood. 2011;118(25):6601–6609. doi:10.1182/blood-2011-07-365189

4. Zaimoku Y, Patel BA, Adams SD, et al. HLA associations, somatic loss of HLA expression, and clinical outcomes in immune aplastic anemia. Blood. 2021;138(26):2799–2809. doi:10.1182/blood.2021012895

5. Savage SA, Viard M, O’hUigin C, et al. Genome-wide Association Study Identifies HLA-DPB1 as a Significant Risk Factor for Severe Aplastic Anemia. Am J Hum Genet. 2020;106(2):264–271. doi:10.1016/j.ajhg.2020.01.004

6. Wang T, Chen Q, Ma Y, et al. Eltrombopag restores T-cell homeostasis in aplastic anemia by regulating oxidative metabolism and reactive oxygen species levels. Ann Hematol. 2026;105(8):323. doi:10.1007/s00277-026-07060-7

7. Single-cell transcriptomics dissects hematopoietic cell destruction and T-cell engagement in aplastic anemia | Blood | American Society of Hematology. Accessed August 8, 2026. https://ashpublications.org/blood/article/138/1/23/475591/Single-cell-transcriptomics-dissects-hematopoietic

8. Ben Hamza A, Welters C, Stadler S, et al. Virus-reactive T cells expanded in aplastic anemia eliminate hematopoietic progenitor cells by molecular mimicry. Blood. 2024;143(14):1365–1378. doi:10.1182/blood.2023023142

9. Single-cell analysis of aplastic anemia reveals a convergence of NK and NK-like CD8+ T cells with a disease-associated TCR signature | Science Translational Medicine. Accessed August 8, 2026. https://www.science.org/doi/10.1126/scitranslmed.adl6758

10. Wu Z, Gao S, Feng X, et al. Human autoimmunity at single cell resolution in aplastic anemia before and after effective immunotherapy. Nat Commun. 2025;16(1):5048. doi:10.1038/s41467-025-60213-6

11. Rosenberg JM, Peters JM, Hughes T, et al. JAK inhibition in a patient with a STAT1 gain-of-function variant reveals STAT1 dysregulation as a common feature of aplastic anemia. Med. 2022;3(1):42–57.e5. doi:10.1016/j.medj.2021.12.003

12. Lin F ching, Karwan M, Saleh B, et al. IFN-γ causes aplastic anemia by altering hematopoietic stem/progenitor cell composition and disrupting lineage differentiation. Blood. 2014;124(25):3699–3708. doi:10.1182/blood-2014-01-549527

13. Dubey S, Shukla P, Nityanand S. Expression of interferon-γ and tumor necrosis factor-α in bone marrow T cells and their levels in bone marrow plasma in patients with aplastic anemia. Ann Hematol. 2005;84(9):572–577. doi:10.1007/s00277-005-1022-8

14. High-dimensional immune profiling using mass cytometry reveals IL-17A-producing γδ T cells as biomarkers in patients with T-cell-activated idiopathic severe aplastic anemia - ScienceDirect. Accessed August 8, 2026. https://www.sciencedirect.com/science/article/abs/pii/S1567576923014893?via%3Dihub

15. Giudice V, Feng X, Lin Z, et al. Deep sequencing and flow cytometric characterization of expanded effector memory CD8+CD57+ T cells frequently reveals T-cell receptor Vβ oligoclonality and CDR3 homology in acquired aplastic anemia. Haematologica. 2018;103(5):759–769. doi:10.3324/haematol.2017.176701

16. Zeng W, Kajigaya S, Chen G, Risitano AM, Nunez O, Young NS. Transcript profile of CD4+ and CD8+ T cells from the bone marrow of acquired aplastic anemia patients. Exp Hematol. 2004;32(9):806–814. doi:10.1016/j.exphem.2004.06.004

17. Olson TS, Frost BF, Duke JL, et al. Pathogenicity and impact of HLA class I alleles in aplastic anemia patients of different ethnicities. JCI Insight. 2022;7(22). doi:10.1172/jci.insight.163040

18. Yoshida M, Sahoo SS, Arnold PY, et al. High-resolution single-cell mapping of clonal hematopoiesis and structural variation in aplastic anemia. Nat Genet. 2026;58(5):1073–1086. doi:10.1038/s41588-026-02587-x

19. Mechanism of Transcriptional Regulation by Methyl-CpG Binding Protein MBD1: Molecular and Cellular Biology: Vol 20 , No 14 - Get Access. Accessed August 8, 2026. https://www.tandfonline.com/doi/full/10.1128/MCB.20.14.5107-5118.2000

20. Fujita N, Watanabe S, Ichimura T, et al. Methyl-CpG Binding Domain 1 (MBD1) Interacts with the Suv39h1-HP1 Heterochromatic Complex for DNA Methylation-based Transcriptional Repression *. J Biol Chem. 2003;278(26):24132–24138. doi:10.1074/jbc.M302283200

21. Chen HTT, Joshi P, Cathelin S, et al. Aberrant splicing of MBD1 reshapes the epigenome to drive convergent myeloerythroid defects in MDS. Blood. Published online June 18, 2026:blood.2025030731. doi:10.1182/blood.2025030731

22. DNA Methylation Represses IFN-γ–Induced and Signal Transducer and Activator of Transcription 1–Mediated IFN Regulatory Factor 8 Activation in Colon Carcinoma Cells | Molecular Cancer Research | American Association for Cancer Research. Accessed August 8, 2026. https://aacrjournals.org/mcr/article/6/12/1841/90184/DNA-Methylation-Represses-IFN-Induced-and-Signal

23. Solomou EE, Keyvanfar K, Young NS. T-bet, a Th1 transcription factor, is up-regulated in T cells from patients with aplastic anemia. Blood. 2006;107(10):3983–3991. doi:10.1182/blood-2005-10-4201

24. Runx3 and T-box proteins cooperate to establish the transcriptional program of effector CTLs | Journal of Experimental Medicine | Rockefeller University Press. Accessed August 8, 2026. https://rupress.org/jem/article/206/1/51/54202/Runx3-and-T-box-proteins-cooperate-to-establish

25. IFN-γ-mediated hematopoietic cell destruction in murine models of immune-mediated bone marrow failure | Blood | American Society of Hematology. Accessed August 8, 2026. https://ashpublications.org/blood/article/126/24/2621/34717/IFN-mediated-hematopoietic-cell-destruction-in

26. Kim MV, Ouyang W, Liao W, Zhang MQ, Li MO. The Transcription Factor Foxo1 Controls Central-Memory CD8+ T Cell Responses to Infection. Immunity. 2013;39(2):286–297. doi:10.1016/j.immuni.2013.07.013

27. Kato H, Itoh-Nakadai A, Matsumoto M, et al. Infection perturbs Bach2- and Bach1-dependent erythroid lineage ‘choice’ to cause anemia. Nat Immunol. 2018;19(10):1059–1070. doi:10.1038/s41590-018-0202-3

28. Zhou X, Xue HH. Cutting Edge: Generation of Memory Precursors and Functional Memory CD8+ T Cells Depends on T Cell Factor-1 and Lymphoid Enhancer-Binding Factor-1. J Immunol. 2012;189(6):2722–2726. doi:10.4049/jimmunol.1201150

29. Roychoudhuri R, Clever D, Li P, et al. BACH2 regulates CD8+ T cell differentiation by controlling access of AP-1 factors to enhancers. Nat Immunol. 2016;17(7):851–860. doi:10.1038/ni.3441

30. Intlekofer AM, Takemoto N, Wherry EJ, et al. Effector and memory CD8+ T cell fate coupled by T-bet and eomesodermin. Nat Immunol. 2005;6(12):1236–1244. doi:10.1038/ni1268

31. Tsao HW, Kaminski J, Kurachi M, et al. Batf-mediated epigenetic control of effector CD8+ T cell differentiation. Sci Immunol. 2022;7(68):eabi4919. doi:10.1126/sciimmunol.abi4919

32. Omilusik KD, Best JA, Yu B, et al. Transcriptional repressor ZEB2 promotes terminal differentiation of CD8+ effector and memory T cell populations during infection. J Exp Med. 2015;212(12):2027–2039. doi:10.1084/jem.20150194

33. Tsukumo S ichi, Unno M, Muto A, et al. Bach2 maintains T cells in a naive state by suppressing effector memory-related genes. Proc Natl Acad Sci. 2013;110(26):10735–10740. doi:10.1073/pnas.1306691110

34. Willinger T, Freeman T, Herbert M, Hasegawa H, McMichael AJ, Callan MFC. Human Naive CD8 T Cells Down-Regulate Expression of the WNT Pathway Transcription Factors Lymphoid Enhancer Binding Factor 1 and Transcription Factor 7 (T Cell Factor-1) following Antigen Encounter In Vitro and In Vivo. J Immunol. 2006;176(3):1439–1446. doi:10.4049/jimmunol.176.3.1439

35. AP-1 activity induced by co-stimulation is required for chromatin opening during T cell activation | Journal of Experimental Medicine | Rockefeller University Press. Accessed August 8, 2026. https://rupress.org/jem/article/217/1/e20182009/132593/AP-1-activity-induced-by-co-stimulation-is

36. A molecular threshold for effector CD8+ T cell differentiation controlled by transcription factors Blimp-1 and T-bet | Nature Immunology. Accessed August 8, 2026. https://www.nature.com/articles/ni.3410

37. Sloand E, Kim S, Maciejewski JP, Tisdale J, Follmann D, Young NS. Intracellular interferon-γ in circulating and marrow T cells detected by flow cytometry and the response to immunosuppressive therapy in patients with aplastic anemia. Blood. 2002;100(4):1185–1191. doi:10.1182/blood-2002-01-0035

38. Sharma V, Kumar P, Kumar R, et al. Interferon-gamma and perforin-positive T cells in acquired aplastic anemia: implication in therapeutic response. Clin Exp Immunol. 2022;207(3):272–278. doi:10.1093/cei/uxab006

39. Single-cell transcriptomic analysis of PB and BM NK cells from severe aplastic anaemia patients - Liu - 2022 - Clinical and Translational Medicine - Wiley Online Library. Accessed August 8, 2026. https://onlinelibrary.wiley.com/doi/10.1002/ctm2.1092

40. Kurki MI, Karjalainen J, Palta P, et al. FinnGen provides genetic insights from a well- phenotyped isolated population. Nature. 2023;613(7944):508–518. doi:10.1038/s41586-022-05473-8

41. GWAS Catalog. Accessed August 8, 2026. https://www.ebi.ac.uk/gwas/studies/GCST90473132

42. GWAS Catalog. Accessed August 8, 2026. https://www.ebi.ac.uk/gwas/studies/GCST90475783

43. Stuart T, Butler A, Hoffman P, et al. Comprehensive Integration of Single-Cell Data. Cell. 2019;177(7):1888–1902.e21. doi:10.1016/j.cell.2019.05.031

44. Fast, sensitive and accurate integration of single-cell data with Harmony | Nature Methods. Accessed August 8, 2026. https://www.nature.com/articles/s41592-019-0619-0

45. Integrated analysis of multimodal single-cell data: Cell. Accessed August 8, 2026. https://www.cell.com/cell/fulltext/S0092-8674(21)00583-3?_returnURL=https%3A%2F%2Flinkinghub.elsevier.com%2Fretrieve%2Fpii%2FS0092867421005833%3Fshowall%3Dtrue

46. UCell: Robust and scalable single-cell gene signature scoring | Computational and Structural Biotechnology Journal. Accessed August 8, 2026. https://spj.science.org/doi/10.1016/j.csbj.2021.06.043

47. Yerly L, Andreatta M, Garnica J, et al. Wounding triggers invasive progression in human basal cell carcinoma. *bioRxiv*. Preprint posted online February 21, 2025:2024.05.31.596823. doi:10.1101/2024.05.31.596823

48. Liberzon A, Birger C, Thorvaldsdóttir H, Ghandi M, Mesirov JP, Tamayo P. The Molecular Signatures Database Hallmark Gene Set Collection. Cell Syst. 2015;1(6):417–425. doi:10.1016/j.cels.2015.12.004

49. Robinson MD, McCarthy DJ, Smyth GK. edgeR: a Bioconductor package for differential expression analysis of digital gene expression data. Bioinformatics. 2010;26(1):139–140. doi:10.1093/bioinformatics/btp616

50. clusterProfiler 4.0: A universal enrichment tool for interpreting omics data: The Innovation. Accessed August 8, 2026. https://www.cell.com/the-innovation/fulltext/S2666-6758(21)00066-7?_returnURL=https%3A%2F%2Flinkinghub.elsevier.com%2Fretrieve%2Fpii%2FS2666675821000667%3Fshowall%3Dtrue

51. ArchR is a scalable software package for integrative single-cell chromatin accessibility analysis | Nature Genetics. Accessed August 8, 2026. https://www.nature.com/articles/s41588-021-00790-6

52. Model-based Analysis of ChIP-Seq (MACS) | Genome Biology | Springer Nature Link. Accessed August 8, 2026. https://link.springer.com/article/10.1186/gb-2008-9-9-r137

53. Weirauch MT, Yang A, Albu M, et al. Determination and Inference of Eukaryotic Transcription Factor Sequence Specificity. Cell. 2014;158(6):1431–1443. doi:10.1016/j.cell.2014.08.009

54. Schep AN, Wu B, Buenrostro JD, Greenleaf WJ. chromVAR: inferring transcription-factor- associated accessibility from single-cell epigenomic data. Nat Methods. 2017;14(10):975–978. doi:10.1038/nmeth.4401

55. STRING database in 2023: protein–protein association networks and functional enrichment analyses for any sequenced genome of interest | Nucleic Acids Research | Oxford Academic. Accessed August 8, 2026. https://academic.oup.com/nar/article/51/D1/D638/6825349

56. Willer CJ, Li Y, Abecasis GR. METAL: fast and efficient meta-analysis of genomewide association scans. Bioinformatics. 2010;26(17):2190–2191. doi:10.1093/bioinformatics/btq340

57. Wang G, Sarkar A, Carbonetto P, Stephens M. A Simple New Approach to Variable Selection in Regression, with Application to Genetic Fine Mapping. J R Stat Soc Ser B Stat Methodol. 2020;82(5):1273–1300. doi:10.1111/rssb.12388

58. Auton A, Abecasis GR, Altshuler DM, et al. A global reference for human genetic variation. Nature. 2015;526(7571):68–74. doi:10.1038/nature15393

59. Bulik-Sullivan BK, Loh PR, Finucane HK, et al. LD Score regression distinguishes confounding from polygenicity in genome-wide association studies. Nat Genet. 2015;47(3):291–295. doi:10.1038/ng.3211

60. Finucane HK, Bulik-Sullivan B, Gusev A, et al. Partitioning heritability by functional annotation using genome-wide association summary statistics. Nat Genet. 2015;47(11):1228–1235. doi:10.1038/ng.3404

